# Medical termination of pregnancy from day 85 to day 153 of gestation: A randomized comparison between administration of the initial dose of misoprostol at home or in the clinic

**DOI:** 10.1101/2024.08.08.24311688

**Authors:** Kristina Gemzell Danielsson, Johanna Rydelius

## Abstract

**Study Protocol:** *Objectives:* The objective of this study is to compare two groups of women requiring termination of pregnancy from the gestational age of 85 days. All women will receive Mifepristone during their first visit to the out-patient ward. One group of women will receive Misoprostol to administer the first dose (vaginally) at home and 2 hours later they will be admitted to the in-patient ward. They will be informed to present earlier if they start bleeding or experience pain corresponding to more than normal menstrual cramping. The other group will receive the first dose of Misoprostol (vaginally) when admitted in the in-patient ward (usually in the morning) according to current practice.

*Primary objective:* To investigate the proportion of women who are successfully treated as day-care patients (day-care being defined as 9 hours from time of admission).

*Design:* The study will be a randomized, controlled, multi-centred open label trial with women recruited from four hospitals in Sweden.

## 3. Synopsis

|  |  |
| --- | --- |
| Study Title | Medical termination of pregnancy from day 85 to day 153 of gestation: A randomized comparison between administration of the initial dose of misoprostol at home or in the clinic |
| ClinicalTrial registration number<br>EuraCT number<br>Sponsor's protocol number | WP2018.PRIMA <u>2</u>.0 |
| Clinical study phase | Phase IV |
| Study objective | To investigate the proportion of women who are treated as day-care patients (day-care being defined as 9 hours from time of admission) while performing a medical abortion from day 85 to day 153 of gestation. |
| Intervention arm | Home administration of 0.8 mg misoprostol pv followed by hospital admission prior to subsequent administration of misoprostol and completion of the abortion |
| Reference arm | Hospital administration of 0.8 mg misoprostol pv followed by subsequent administration of misoprostol and completion of the abortion |
| Study Design | A prospective, randomized two armed parallel open label trial. |
| Main patient inclusion criteria | 1. Women 18 years of age or above eligible for medical abortion<br>2. The pregnancy is single and intrauterine<br>3. Willing and able to participate after the study has been explained |
|  | 4. Signed informed consent |
| Main patient exclusion criteria | 1. Allergy towards mifepristone or misoprostol<br>2. A medical condition or disease that requires special treatment, care or precaution (e.g. corticosteroid or anticoagulant therapy) in conjunction with abortion<br>3. IUFD/missed abortion<br>4. Foetal malformation that could be expected to affect the induction-to-abortion interval<br>5. Duplex pregnancy<br>6. Application not approved by the Board of Health and Welfare (from gestational week 18+1) |
| Number of patients | 896 women |
| Primary variable | Complete abortion as day care patient (9 h from admission to the out patient clinic). No overnight hospitalization required. |
| Statistical analysis | Patient flow during trial will follow the CONSORT chart. For demographic variables the full analysis set, FAS (all subjects randomized), will be presented with descriptive statistics. All subjects randomized having known primary outcome (time for admission, and discharge) will be included in the main analysis in their randomized groups. If attempts fail to locate and contact a woman lost to follow-up, and her final outcome of treatment is not known that woman will be excluded from the main efficacy analysis.<br>Crude rates for each outcome will be calculated in each arm and exact confidence intervals calculated. |
| Study duration | 2017 Q4-2018 Q1/2 approvals<br>Q2 2018 Trial start. Recruitment of patients will start Q2 2018 and is estimated to be concluded within two years. Time between the last patient recruited and the last patient last visit /= telephone follow-up) (LPLV): will be 4 weeks. Data analysis and presenting |
|  | will take another 6 months. |

## 4. Overall aim

The overall aim is to optimize the protocol for women requiring a late medical termination of pregnancy; that is from day 85 to day 153 of gestation. The aim is to explore if possible to shorten the duration of stay in hospital and to increase the number of women who perform a late medical abortion as a day-care procedure.

## 5. Benefit risk assessment

Participation will be voluntary by healthy well informed women. Theoretically, following administration of mifepristone and in addition starting the first dose of misoprostol administration at home, there is a risk of aborting at home or on the way to the hospital, So far this has rarely been reported (3, 6, 9, 11). The clinical routine of administering the first dose of misoprostol is already in use in some of the largest abortion clinics in Sweden. An evaluation of this practise is therefore appropriate.

Uterine rupture in medical second trimester pregnancy is rare and occurs typically when oxytocin is given despite an unripe rigid and closed cervix. Rupture doesn’t occur without uterine contractions. Rupture following the first dose of misoprostol has to our knowledge not been reported.

A systematic review of uterine rupture with misoprostol induced second trimester abortion further reported the risk to be 0.28% among women with a prior cesarean section and 0.04% among women without a prior C-section (12). Information on uterine rupture following the mifepristone-misoprostol regimen in second trimester abortions is limited. Theoretically it is likely that pretreatment with mifepristone reduces the risk for uterine rupture due to improved cervical ripening prior to onset of contractions and due to lower total doses of misoprostol needed for expulsion.

Data on uterine rupture is generally scarce and summarized in Lalitkumar et al,(3).

### 5.1 Risk mitigation plan

For safety reasons no patient should be alone at home when taking the first dose of misoprostol. Women who have already developed regular contractions should not administer misoprostol at home.

## 6. Scientific background

Abortion is defined as termination of pregnancy before the foetus is viable outside the uterus. The second trimester is a period ranging from 13 to 28 weeks of gestation. Second trimester abortions constitute only 10-15 % of all abortions worldwide, but they are responsible for two-thirds of major abortion-related complications. The number of second trimester abortions is believed to gradually increase due to the wide-scale introduction of prenatal screening programs globally. In this study a late abortion is defined as a termination of pregnancy occurring from 12 gestational weeks (GW) and 1 day (85 days) to 21 GW and 6 days (135 days), which is the legal limit in Sweden. During 2016, 38 000 abortions were reported in Sweden and 6, 4 % were performed after 85 days (1 – 3).

Traditionally dilatation and evacuation has been the standard method to perform a second trimester abortion, and still is in many parts of the world. But even when dilatation and evacuation of the uterus is performed by a skilled gynaecologist there is a risk of serious complications such as uterine perforation and cervical laceration. Guidelines in Sweden therefore recommend a medically induced termination of pregnancy (TOP) after 85 days. Even though medically induced second trimester TOP has proven to be a safe method the risk of surgical evacuation and infection are increased compared to first trimester medical TOP. In Sweden medically induced abortion is performed in 91, 6 % of the total number of abortions (both early and late), and in 98, 3% of the late abortions (2016) (1 – 4).

Medical abortion with Mifepristone followed by a prostaglandin analogue is the current method of choice and has been shown to be safe and effective. The most commonly used combination of drugs is Mifepristone and Misoprostol. Mifepristone is a synthetic steroid which acts as an antiprogestin. Treatment with Mifepristone softens the cervix and sensitizes the pregnant uterus to exogenous prostaglandin. Misoprostol is a prostaglandin E1 analogue that induces cervical ripening and stimulation of myometrial activity which lead to expulsion of the pregnancy (2, 3, 5 – 8).

It has been well proven that pre-treatment with Mifepristone 36 – 48 hours before administration of Misoprostol increases the success rate and shortens the induction-to-abortion interval (time interval from the first tablet of Misoprostol to foetal expulsion) and reduces the amount of Misoprostol tablets used for second trimester abortion with a median induction-to-abortion interval of 6 – 6, 5 hours. Most women require multiple doses before the abortion is completed, with a median of 3 doses. The combined regime with Mifepristone and Misoprostol has a success rate as high as 97%, for second trimester abortions (3, 5 – 9).

The medical protocol covering the combined regime is described in detail below. In short, a tablet with 200 mg of Mifepristone is administered orally after a policlinic visit with clinical examination and estimation of the gestational age. Thirty-six to 48 hours later the woman returns to the clinic and is admitted to the in-patient ward where she administers 0, 8 mg of Misoprostol vaginally. If the woman does not abort within 3 hours she will receive 0, 4 mg of Misoprostol every 3^rd^ hour vaginally or orally/sublingually to a maximum of 4 times, until foetal expulsion (3, 10).

Surgical evacuation of the uterus is not routinely performed following a late medical abortion. It should only be undertaken if there is clinical evidence that the abortion is incomplete; for example if the placenta is not expelled or if a profuse bleeding occurs. In a case series of second trimester medical abortions, only 2, 5 – 11 % of women needed surgical evacuation. After the abortion is completed the women should be observed in the hospital to monitor the vital signs and the amount of vaginal bleeding (3, 8, 9). However, guidance on the duration of this post-abortion observation is lacking.

Although considered a safe procedure, medical abortion during the second trimester has several limitations including the need for hospitalization. Studies have shown that 60 – 80 % of second trimester abortions receiving Misoprostol, following administration of Mifepristone, aborted within 8 – 12 h, confirming that the majority of late abortions can be managed as day-care procedures. In a review from 2004 covering 1002 cases of mid-trimester medical abortions 72, 4 % was completed on a day-care basis. Despite the relatively high ratio of day-care treatment it still seems like a certain proportion of women undergoing a late medical abortion will require inpatient care for medical, social or geographical reasons. There are currently no available national data of the number of patients in Sweden requiring an over-night stay after performing a late medical abortion (3, 5, 9).

Treating second trimester abortions on a day-care basis has several advantages. One study on outpatient second trimester abortions showed that women randomized to the outpatient arm favoured the option because of the privacy it allowed. Being at home for at least part of the process allowed extended contact and support from the immediate family. Another benefit of an ambulatory second trimester termination includes a potential decreased risk of nosocomial infection (11).

Since first trimester medical abortions performed at home have shown to have positive economic aspects for patients as well as for the health care system it could be assumed that day-care for second trimester abortions is the more cost-effective method as well.

Considering the positive aspects of late termination of pregnancy as a day-care procedure it is important to further improve the medical protocol to shorten the duration of time in hospital so that more women can be managed as day-care patients.

## 7. Objectives

The objective of this study is to compare two groups of women requiring termination of pregnancy from the gestational age of 85 days. All women will receive Mifepristone during their first visit to the out-patient ward. One group of women will receive Misoprostol to administer the first dose (vaginally) at home and 2 hours later they will be admitted to the in-patient ward. They will be informed to present earlier if they start bleeding or experience pain corresponding to more than normal menstrual cramping.

The other group will receive the first dose of Misoprostol (vaginally) when admitted in the in-patient ward (usually in the morning) according to current practice.

### 7.1 Primary objective

To investigate the proportion of women who are successfully treated as day-care patients (day-care being defined as 9 hours from time of admission).

### 7.2 Secondary objectives

The two groups will also be compared regarding difference in length of stay in the hospital (h) (from admittance to the in-patient unit until discharge) and success rate at 24 h (success will be defined as the expulsion of the foetus), difference in the induction-to-abortion interval (defined as the time from the first dose of Misoprostol until the expulsion of the foetus) (mean/median), doses of Misoprostol used (mean/median), frequency of complications (direct and up to 4 weeks), surgical interventions needed (such as evacuation rates) and side effects experienced. Patient satisfaction will be compared between the groups using pretested questions.

## 8. Design

The study will be a randomized, controlled, multi-centred open label trial with women recruited from four hospitals in Sweden.

## 9. Setting/centres

The Department of Gynecology and Reproductive medicine at Östra sjukhuset, Sahlgrenska University Hospital, Gothenburg.

The Department of Obstetrics and Gynaecology, Karolinska University Hospital, Stockholm.

The Department of Obstetrics/Gynaecology, Stockholm South General Hospital (Södersjukhuset), Stockholm.

The Department of Obstetrics and Gynaecology, Danderyd hospital, Stockholm.

## 10. Participants

Women requesting a termination of pregnancy with live pregnancies from day 85 to day 153 of gestation (based on ultrasound). Reasons for termination of pregnancy being social, medical or foetal indications (such as malformations which are not expected to affect time to expulsion, see below).

### 10.1 Interventions and controls

Women requesting a termination of pregnancy (at 85 – 153 days of gestation) will be randomized into two groups:

Group 1) Administration of 4 tablets of 0,2 mg of Misoprostol vaginally by the patient herself at home before presenting in the inpatient unit 2 hours later.

Group 2) Administration of 4 tablets of 0,2 mg of Misoprostol vaginally after admission to the inpatient unit.

After administration of the first dose of Misoprostol women in both groups will be treated according to the clinical routine protocol (see below). A follow-up will be undertaken two to four weeks after discharge.

## 11. Outcome measures

### 11.1 Main outcome measure

The proportion of women who are successfully treated as day-care patients will be the main outcome (day-care being defined as 9 hours from time of admission).

### 11.2 Secondary outcome measures

Secondary outcome measures will include the difference in time spent in hospital (h), the difference in the induction-to-abortion interval (defined as the time (min) from the first dose of Misoprostol until the expulsion of the foetus) (mean/median), the success rate of the termination of pregnancy at 24 hours (success will be defined as the expulsion of the foetus), and doses of Misoprostol used (mean/median). The difference in frequency of complications (during treatment and until the follow-up visit), the difference regarding surgical interventions for incomplete or retained placenta (evacuation rates) and side effects will be measured. Other outcome measures will be time for and grade of maximal pain during the abortion and maximal pain measured on a VAS (visual analogue scale) (before and 30 minutes following administration of pain treatment) and doses of pain medication given. Acceptability with allocated treatment (evaluated by two pretested questions) and the difference in number of women who need to be admitted to the hospital earlier prior to planned admission and for which reasons will be registered.

## 12. Eligibility criteria

### 12.1 Inclusion criteria

Women aged >/= 18 years requesting a termination of pregnancy, for social, medical or foetal indications and the gestational age being determined to 85 – 153 days (with ultrasonography), willing and able to understand and participate after the study has been explained, with good understanding of Swedish or English language, in general good health, with a single intra-uterine pregnancy, who have given their informed consent will be eligible for the study.

### 12.2 Exclusion criteria

Women who do not wish to participate or who are unable to communicate in Swedish or English. Women carrying a non-viable pregnancy (confirmed by ultrasonography).

Women with a history or evidence of disorders that represent a contraindication to the use of Mifepristone or Misoprostol (adrenal pathology, steroid dependent cancer, porphyria, known allergy to the medication).

Women with pre-existing health conditions for whom the execution of a medical abortion as judged by the investigator could compromise their condition will also be excluded. Examples of health conditions of concern are: severe anaemia, severe asthma, haemorrhagic disorders, treatment with anticoagulants or corticoids, severe cardiac disease or hypertension, severe liver disease, severe kidney disease or severe psychiatric disorders.

Fetal malformation that may impact time–to-expulsion (such as hydrocephalus, hydrops/edema) or a duplex pregnancy.

## 13. Trial process and data collection

The study protocol is designed according to the CONSORT recommendations. The study is a randomized and controlled open label trial. The study flow is planned as follows:

### 13.1 Enrolment

All eligible women, requesting a termination of pregnancy, with live pregnancies from day 85 to day 153 of gestation, will be invited to participate in the study at the initial outpatient consultation. The women will receive detailed oral and written information and have the possibility to ask questions regarding the study according to ICH-GCP. Written informed consent will be signed by the attending physician and the women before randomization.

### 13.2 Allocation and treatment

Designated study nurses and doctors will be responsible for recruiting and examining study participants at the outpatient clinic. A medical history will be obtained to identify contraindications and risk factors or complications to the pregnancy. The history of the woman will include personal and family history of relevant diseases, current use of medications, allergies, obstetric and gynaecological history or any bleeding tendencies.

Physical examination will be performed including vaginal examination and ultrasonography to establish the gestational age. All women will be screened for bacterial vaginosis and Chlamydia infection and treated if positive. Vital signs (pulse, blood pressure), length, weight, haemoglobin, blood group and Rhesus typing will be documented by the assisting nurse or midwife.

Eligible women with a pregnancy from day 85 to day 153 of gestation who wish to participate in the study and who sign an informed consent will be randomized into either Group 1 or 2:

After randomization, all women will swallow Mifepristone 200 mg at the outpatient clinic. The day when Mifepristone is administered counts as Day 1. The woman will receive the date and time when she shall return to the inpatient unit to continue the treatment. The time interval between the administration of Mifepristone and Misoprostol is 36-48 hours according to national guidelines.

On Day 3 the woman will administer Misoprostol:

Group 1) Administration of 4 tablets of 0, 2 mg of Misoprostol vaginally by the patient herself at home before admission to the inpatient unit two hours later.

or

Group 2) Administration of 4 tablets of 0, 2 mg of Misoprostol vaginally after arrival to the inpatient unit.

When admitted to the inpatient unit the women will follow the clinical protocol which means that if she does not abort within 3 hours of the first vaginal dose of Misoprostol she will receive a second dose of 2 tablets of 0,2 mg Misoprostol sublingually and then every 3^rd^ hour until expulsion of the foetus (maximum of 4 doses).

Pain medication will be administered as NSAIDs and Paracetamol which will be administered every eight hours. Oxikodon 5 mg (orally) and/or Morphine 5 mg (injectable) will be administered as required. Other analgesic options are transcutaneous electrical nerve stimulation (TENS), paracervical blockade (PCB) with Carbocain-Adrenalin or epidural anaesthesia.

After the abortion the women should be observed in the hospital for a minimum of 1 hour to monitor the vital signs and the amount of vaginal bleeding. Reasons for staying longer than the clinical routine will be documented. Before being discharged the woman will be asked regarding pain, satisfaction with the treatment and experienced side effects.

After being discharged the woman is recommended to contact the clinic if heavy vaginal bleeding, severe abdominal pain (where prescribed analgesic do not reduce the pain), malaise or fever >38 °C should occur. The woman will be booked for a follow-up with a midwife 2 - 4 weeks after discharge from the ward. Since previous studies have shown that the loss for the follow-up visit may be as high as 20 %, the follow-up could also be conducted by telephone-call or e-mail to reach as many women as possible. The follow-up a will cover contraceptive plan (possible insertion of an IUD (intra-uterine device)) or birth control implant (if not already done) and any extra visits or complications since discharge. If the reason for termination of pregnancy was a foetal malformation, the patient will also be booked to the referring obstetrician.

The initial judgment about the outcome of medical abortion is made at the clinic before discharge home. The outcome is classified on the basis of the woman’s history and the clinical findings, as one of the following:

1. Complete abortion - no additional treatment needed for foetal and complete placental expulsion - uterus empty
2. Incomplete abortion - vacuum aspiration or exeresis done due to remnants of tissue in uterus

The final outcome of the medical abortion will be assessed at the follow-up

### 13.3 Randomization procedure

The randomization ratio between intervention and control group will be 1:1 in permuted blocks of 4-12. Randomization will be performed by the midwife in the abortion clinic at the time of the first visit after the patient has been judged eligible for participation and has signed informed consent in the presence of the attending physician. Allocation to intervention or control will be performed via randomization through a study specific internet allocation site after an online checklist of inclusion and exclusion criteria has been completed. All women will be identified through a patient log with name and personal identification number and the randomization number (patient study ID). The randomization number will be used on all paper and eCRFs (case record form), for future identification. The study will for practical and ethical reasons not be blinded. The medical abortion will be performed according to the Swedish national guidelines.

### 13.4 Discontinuation

After recruitment, women may withdraw from the trial at any time without giving any reason if they do not wish to participate. If interim analysis shows that a higher rate of women (compared to what is previously known, up to 4%) included in the study will abort at home or on the way to the hospital the study will be interrupted as judged by the DSMB (11).

### 13.5 Discontinuation of the study itself

The regimen for medical abortion that will be used in the study is well known. Medical abortion when started can not be interrupted since no other abortion methods are available for second trimester abortions in Sweden. If a woman regret the decision to terminate the pregnancy she can interrupt the abortion, and the local clinical protocol will be used. The proposed regimen is the one recommended by SFOG (Svensk Förening för Obstetric och Gynekologi). Thus a discontinuation of the study is not anticipated based on any known incidence of SAE or any other reasons.

## 14. Registration and reporting of suspected adverse events

Complications and the number of women requiring surgical intervention will be recorded. Any unexpected and serious side effect or suspected adverse reaction will be immediately reported to the project principal investigator, the sponsor as well as to the trial steering committee. The regional ethics committee will also be notified according to local regulations.

Any suspected unexpected serious adverse event that occurs during the study will be reported to the principal investigator, the sponsor immediately on notification. This will also be recorded in the patient file which will be kept (with the other study documents) for at least 15 years.

### 14.1 Adverse events

Definition: An adverse event (AE) is defined as any untoward medical occurrence in a patient (or clinical investigation subject) administered a pharmaceutical product and which does not necessarily have to have a causal relationship with this treatment. An AE can therefore be any unfavourable and unintended sign (including abnormal lab finding), symptom, or disease temporarily associated with the use of a medical product, whether or not considered related to this medical product.

A Serious AE (SAE) is defined as any event that:

- results in death
- is life threatening
- results in hospitalization or prolonged stay in hospital
- results in temporary or permanent morbidity
- results in congenital malformation/teratogenic effect
- Any other serious event

All adverse events will be reported by staff in the patient chart or self-reported by the patient in the questionnaires. All reported adverse events will be recorded in the patient records and scrutinized by the investigator and monitor to judge the severity (mild, moderate, severe) and relation to treatment (not related, possibly related, probably related).

All serious adverse events will be reported by the sponsor to the Medical Products Agency on paper for subsequent registration into the EudraVigilance database.

### 14.2 Reporting of AE, SAE and SUSARs

- All patients will be asked for adverse events at every visit/contact with the investigator or study nurse/midwife with the exception for what is mentioned under 13.1
- Adverse events will be reported irrespective if the event has any relation with the study drug or not
- All Adverse events must be recorded in the CRF, defining the relationship to study medication, severity and seriousness. Adverse Events should also be recorded in the patient records if applicable
- If a reported SAE is present when the patient completes the study the event must be followed for up to three months past the final visit or followed until final outcome is known or the condition is stable
- AE/SAE will be registered until the end of the study

### 14.3 Reporting of SAE and SADE and USADE

Any SAE that might have led to a SAE if: a) suitable action had not been taken or b) intervention had not been made or c) if circumstances had been less fortunate and in addition new findings/updates in relation to already reported events must be reported to the sponsor on a separate SAE report form within 24 hours of the initial notification of the event-for device deficiencies regulatory requirement is within 3 days-however, a 24 hour period will be maintained for all serious adverse events in this trial.

- The initial notification to the sponsor can be made on a SAE-report form, by e-mail or telephone.
- At the time of initial reporting the investigator must provide a minimum requirement, the patient number, birth date, description of the SAE and a preliminary assessment of causality.
- Supplemental information shall be reported by the investigator to the sponsor as soon as possible. If the SAE is fatal or life-threatening the investigator is responsible to report follow-up information to the sponsor within 5 days after the initial report
- For all reportable events as described above which indicate an imminent risk of death, serious injury, or serious illness and that requires prompt remedial action for other patients/subjects, users or other persons or a new finding to it: immediately, but not later than 2 calendar days after awareness by sponsor of a new reportable event or of new information in relation with an already reported event.
- Any other reportable events as described above or a new finding/update to it: immediately, but not later than 7 calendar days following the date of awareness by the sponsor of the new reportable event or of new information in relation with an already reported event.

### 14.5 Reporting SUSAR

#### Sponsor is obliged to report

- Any suspected serious unexpected reaction of the study drug (SUSAR) which is life threatening or fatal to the Medical Products Agency MPA (“Läkemedelsverket, LMV”) as soon as possible and not later than 7 days after the responsible investigator has been aware of the event. The report should be sent to MPA and the Regional Ethical Review Board (RERB). Relevant follow-up information of the report should be sent within additional 8 days (maximum of 15 days from initial report).
- A suspected serious unexpected reaction of the study drug/device (SUSAR) that is **not** life threatening or fatal should be reported as soon as possible but within a maximum of 15 days to the MPA and the RERB.
- All SUSARs should be reported to the MPA on a CIOMS-form and with assistance of the MPA also reported into the Eudra Vigilance Clinical Trial Module

During the study, the sponsor should report all SAEs with a suspected relation to the study drug, and SUSAR, to the MPA and RERB once a year. Moreover, a safety report concerning the patients included in the study should be supplied together with a summary on a risk/benefit evaluation for the patients participating in the study. The sponsor is obliged to inform all investigators about occurring SUSARs during the study period.

### 14.6 Annual Safety Report

The sponsor will assemble the annual safety report according to the instruction of the Medical Product Agency.

## 15. Trial start date

It is estimated that ethics permission will be granted in Q4 2017. Recruitment of patients will start Q2 2018 and is estimated to be concluded within two years. Time between the last patient recruited and the last patient last visit /= telephone follow-up) (LPLV): will be 4 weeks. Data analysis and presenting will take another 6 months.

## 16. Statistical analysis; power calculations – sample size

### 16.1 Sample size

The main outcome measure will be the difference in the proportion (%) of women who are treated as day care patients between the two groups. A difference of 10 percent could be regarded as clinically significant.

784 patients (392 per group) are required to have a 90% chance of detecting, as significant at the 5% level (2-sided test), an increase in the primary outcome measure from 70% in the control group to 80% in the experimental group. To compensate for loss to follow-up 896 women will be recruited.

Patient flow during trial will follow the CONSORT chart. For demographic variables the full analysis set, FAS (all subjects randomized), will be presented with descriptive statistics. All subjects randomized having known primary outcome (day care treatment) will be included in the main efficacy analysis in their randomized groups. If attempts fail to locate and contact a woman lost to follow-up, and her final outcome of treatment is not known that woman will be excluded from the main analysis.

Crude rates for each outcome will be calculated in each arm and exact confidence intervals based on the binomial distribution calculated as described in Armitage and Berry (1994).

Comparisons of percentages, such as the difference in number of women being treated as day-care patients, will be calculated by chi^2^-test or Fischers exact test. Confidence intervals for differences between percentages will be calculated. Comparisons of differences in number of doses and cumulated dose of medication used and hours spent at hospital will be analysed by large sample statistical methods. VAS-scale and difference in patient satisfaction will be analysed by non-parametric rank-tests.

Results will be considered statistically significant if P-value < 0,05. Analyses will be carried out using SPSS (24.0) software package.

The groups will be compared regarding maternal age, parity, number of former abortions, gestational age, maternal BMI, smoking and reasons for termination of pregnancy.

## 17. Assessment of ‘induction-to-abortion interval’

Induction-to-abortion interval is the time in hours from the start of misoprostol administration until the expulsion of the foetus. If the woman aborts before misoprostol administration, the time to abortion will be negative. According to previous experience up to two thirds of the expulsions take place in the clinic during the first six- to eight hour-observation period after misoprostol administration (first dose). The time of expulsion will be recorded on the data collection forms.

The main efficacy analysis will include all women randomized for whom outcome is known. Criteria for exclusion from per-protocol analysis are the following:

- essential data are missing from the participant’s records making it impossible to judge treatment outcome (same as for main analysis)
- any violation of the study protocol, including violation of eligibility criteria treatment non-compliance

The sponsor will be overall responsible for data analysis and reporting together with the co-investigators

## 18. Description of the study drug

Misoprostol (product name Cytotec®; tablet 0, 2 mg Pfizer) is a prostaglandin E1 analogue (15-deoxy-16-hydroxy-16-methyl). It induces cervical ripening both by a direct effect on the cervix and by a concomitant stimulation of myometrium activity and leads to expulsion of a pregnancy. Misoprostol can be administered orally, vaginally, sublingually and rectally. The most common adverse effects of Misoprostol are nausea, vomiting and diarrhoea and are characteristics of the prostaglandin stimulatory effect on the gastrointestinal tract. Other common side effects are abdominal pain, chills, shivering and fever. These side effects usually subside or resolve 24 hours after the last dose of Misoprostol. Serious complications such as uterine rupture and heavy bleeding requiring blood transfusion are rare. Infection is not common after medical abortion by Misoprostol and the incidence is estimated to <1% (2, 3, 7, 8, 13).

Mifepristone (product name Mifegyne®; tablet 200 mg Nordic Drugs) is a synthetic steroid which acts as an antiprogestin. It binds with high affinity to progesterone receptors in target cells of the endometrium and myometrium, thus inhibiting the effect of the hormone. Blockage of the progesterone receptor results in vascular damage, decidual necrosis and bleeding. Treatment with Mifepristone will soften the cervix and sensitize the pregnant uterus to exogenous prostaglandin. The sensitivity of the myometrium is increased by 5 times with maximum effect on uterine contractility and cervical ripening 36-48 h following treatment. The benefits of pre-treatment with Mifepristone are well established. Around 0,2-0,4 % of women abort with Mifepristone only (2, 3, 6).

### 18.1 Distribution and handling of the study drug

Misoprostol and mifepristone will be used according to the clinical routine.

The off label use is close to the approved use. Sublingual administration results in very similar effects to vaginal administration of misoprostol but drug absorption is not affected by vaginal bleeding. Vaginal misoprostol is approved as Medabon (SunPharma) but not available in Sweden.

The misoprostol tablets are packed in aluminium blisters. Each participating woman will get the mifepristone tablet to swallow in the abortion clinic after inclusion in the trial (Day 1). For women randomized to Group 1 misoprostol tablets for home use will be provided in the same way and the same dose as for home use in early medical abortion up to 9 weeks of gestation. Additional misoprostol will be provided to the participating woman after admission to the clinic until abortion as needed. Women randomized to Group 2 will receive misoprostol in the clinic.

### 18.2 Justification of the choice of misoprostol in the form of Cytotec instead of Medabon

The recommended dosage of misoprostol for medical abortion beyond 12 weeks and 0 days is a loading dose of 800 microgram of misoprostol administered 36 to 48 hours following mifepristone. This dosage is approved in Sweden for medical abortion up to 9 weeks (Medabon, combi pack) and investigated in Sweden for medical abortion at 9 to 22 weeks of gestation (Concept Foundation, **EudraCT Number : 2013-004294-27**, Dnr 5.1-2013-89810) multicenter trial) with repeat doses of 400 microgram misoprostol (using misoprostol tablets from Medabon combipacks) administered up to expulsion of the pregnancy.

Misoprostol in Medabon has been accepted as being equivalent in bioavailability of misoprostol free acid to misoprostol free acid in Cytotec. Since Medabon is no longer available on the Swedish market Cytotec replace Medabon in clinical practice. This implies that the benefit-risk balance of Cytotec in the study population is positive.

## 19. Applications, and substantial amendments

The study has received RERB approval at Karolinska Institutet with Dnr. 2017/2312-31/2. If substantial changes in the protocol or patient information or consent will be undertaken, an applications will be sent to the MPA using the “Substantial amendment notification form” as well as the RERB. The study will not start until all approvals by the MPA and RERB are in order.

Changes in the protocol that might affect the safety, physical or psychological integrity, change the scientific value of the study or may be considered as important of any other reason should be considered as substantial changes. If the study has been temporarily stopped and then re-started, this should also be considered as a substantial change. The study will be performed according to the protocol. All protocol deviations will be noted and kept in a protocol deviation log describing the deviation and actions undertaken. All protocol deviations will be monitored by the PI and the appointed monitor and classified as minor, major or severe. All severe protocol deviations will be reported to the RERB and the MPA.

## 20. Trial data and safety monitoring committee (DSMB)

The trial will be conducted under the supervision of the principal coordinating investigator and the sponsor who are also responsible for the analysis and writing-up of the results in collaboration with the co-investigators. A research nurse not directly involved in the trial will be responsible for monitoring the study on at least a monthly basis.

Data from hospital records and other data generated from the study will be entered into CRFs stored in a data base. A trial DSMB will be appointed.

The committee will consist of three experienced clinical researchers and specialists in Obstetrics and Gynecology with good knowledge in medical abortion and otherwise not involved in the trial.

Outcome on heavy bleeding requiring blood transfusion, severe pain, expulsion, and AEs following the first dose of misoprostol at home until admission to the clinic will be reviewed and analyzed every three months. If the rate of expulsion at home is above 4 percent or need for blood transfusion is above 1 percent or pain is rated above 8 on the VAS scale or AEs judged as probably/ possibly related to the misoprostol treatment or study conduct exceed those in the clinic group at the same time point (up to 2 hours following the first dose of misoprostol) by more than 20% the study will be interrupted.

The DSMB will have access to anonymous data extracted from the electronic CRFs. A review will be done every three months.

## 21. Study Management

### Monitoring

Before the study starts an external monitor, a research nurse/midwife (trained and familiar in ICH-GCP and independent of study conduct) will perform an initiation visits at each site to ensure that the essential documents and study records are stored and kept safe in Investigator files. Monitor will also discuss the details regarding the randomization procedure and to see to that staff is familiar with the concept of documenting according to ICH-GCP in medical records and in CRF. Monitor will also visit the site during the study regularly to conduct source data verification, and perform a visit after the study has ended to close the site and gather essential documents to sponsor. Upon request, the monitor, sponsor (auditor) and/or sponsor representative, medical authorities are entitled direct access to source data under the condition that there is a valid signed secrecy agreement. Source documents are considered e.g. patients medical records, ultra scan pictures and other relevant documents that contains data that verifies the accurateness and is stated in the approved protocol.

Declaration of End of Trial Notification will be sent to the MPA at the latest 90 days after EOT (or at premature discontinuation of the trial),

## 22. Archive

All study related documents will be kept in archive for at least 10 years after study completion according to national law.

## 23. Ethics committees and other regulatory boards

Permission will be obtained from the regional ethics committees and from The Medical Products Agency in Sweden.

## 24. Ethical issues

Permission has been obtained from the regional ethics committee.

The RCT will be posted at www.clinicaltrials.gov. The trial will be conducted in compliance with the protocol, GCP, Helsinki declaration and the applicable regulatory requirements, including the Swedish legislation.

Women will receive oral and written study specific information and an informed consent will be signed by the participating women and the investigators prior to any participation in the trial. PUL (personuppgiftslagen) will be followed. Confidentiality will be guaranteed. Data recorded on CRFs will be coded and stored with the PI in approved and locked storages. Data will only be presented at statistical group level.

## 25. Ethical considerations

Since termination of pregnancy might be considered a sensitive subject in the general population the investigators must show consideration and guarantee full confidentiality to the study participants. As pointed out above the performance of a medically induced second trimester termination of pregnancy is considered a safe procedure and this study follows guidelines currently in use. The medication (Misoprostol and Mifepristone) has proven to be safe and tolerable drugs with very few contra-indications. Theoretically, following administration of mifepristone and in addition starting the first dose of misoprostol administration at home, there is a risk of aborting at home or on the way to the hospital, but so far this has rarely been reported (3, 6, 9, 11).

Women will be given information on the possible risks of home abortions and will be able to withdraw from participation in the study at any point without having to justify their decision and without any negative impact on their abortion care. The participating subjects will be informed both orally and in written before they give informed consent.

Abortion could be a very personal question and there might be a risk that the woman can feel that her integrity is affected. The subjects will however be informed before the study starts and before they give their consent. Moreover, all data will be coded and at the time of analysis there will be no possibility to link the information to a specific woman.

## 26. Significance

Since more medical interventions are carried out as day care procedures, hence reducing costs, improving care by specialists and increasing patient satisfaction, it is important to further improve the medical guidelines steering the care of women undergoing a late termination of pregnancy. Following extended prenatal screening programs there is also a world-wide increase in demand for second trimester medical abortions. This requires refined protocols and patient-centred care with highest possible patient satisfaction.

## Data Availability

All data produced in the present study are available upon reasonable request to the authors

## 1. Abbreviations

GA: gestational age
GW: gestational week
TOP: termination of pregnancy
VAS: visual analogue scale
CONSORT: Consolidated Standards of Reporting Trials
TENS: transcutaneous electrical nerve stimulation
PCB: paracervical block
IUD: intra-uterine device
CRF: case record form
AE: adverse event
LPLV: last patient last visit
DSMB: Data and Safety Monitoring Board
PUL: Swedish: personuppgiftslagen; personal records act
ICH-GCP: International Council on Harmonisation of Technical Requirements for Pharmaceuticals for Human Use – Guideline for Good Clinical Practice

